# Reliability and validity of physical function tests and ADL survey questions in women living in rural, highland Ethiopia

**DOI:** 10.1101/2023.07.05.23292266

**Authors:** Jenna Golan, Anna Thalacker-Mercer, John Hoddinott

## Abstract

In rural, highland Ethiopia, physical function, which is the physical ability to fulfill one’s daily roles and responsibilities, may be compromised by a lack of access to nutrition, healthcare, and sanitation. Decreased physical function would be detrimental to health and income-generating activities. Unfortunately, there is a lack of validated methods to measure physical function in adult women in this region. This validity study evaluated the feasibility and reliability of physical tests, including the sit-to-stand (STS) and usual gait speed (UGS). The physical tests were used to assess the validity of context-appropriate activities of daily living (ADL) questions. The results of the STS were used to calculate a power index (P_sts_) which accounted for body mass and leg length. Feasibility was ascertained qualitatively. Reliability was assessed by comparing the results of the tests and questions between each visit using either Cohen’s κ or Pearson’s ρ. Validity was assessed by regressing the responses to the ADL questions against P_sts_, controlling for relevant participant characteristics. Study participants consisted of 316 women between the age of 18 and 45 years, living in rural Tigray, Ethiopia, that had previously participated in an impact evaluation of a safety net program. Over a one-week period, participants completed the STS and UGS tests and responded to the ADL questionnaires three times. P_sts_ was determined to be a feasible, reliable, and valid physical function test for women in rural, highland Ethiopia. UGS lacked feasibility and reliability. The validity of the ADLs was inconclusive. The P_sts_ will be an essential tool for improving physical function and, subsequently, health and quality of life in rural Ethiopia.

## Introduction

A person’s ability to functionally, independently, and physically fulfill their activities of daily living (ADL) is known as physical function (1,2). It is both a cause and consequence of body size and composition and physical activity. Intrinsically, physical function is essential for overall health and quality of life (3,4). Instrumentally, physical function can identify people at risk for further declines in health (5–9), requiring additional care (4), or having future health expenditures (10). Additionally, studies have used physical function as an outcome measure to determine the impact of interventions targeting health (11–13). Outside of health measures, physical function impacts social networks (14,15) and income-generating activities (16–18).

Medical practitioners and researchers in high-income countries have extensively measured physical function in a variety of populations, such as older adults with and without dementia (5,19–23) and people with chronic illness or pain (e.g., arthritis) (24–26). However, measuring physical function has been underused in low- and middle-income countries (LMIC) populations, with few exceptions (19,27–29).

Measuring physical function in LMIC is essential because daily life is physically demanding. According to the 2016 Ethiopia Demographic and Health Survey (DHS), of the employed women living in rural Ethiopia, approximately 65% were employed in skilled manual labor, unskilled manual labor, or agriculture. Additionally, 53% of women in rural households spent more than 30 minutes fetching drinking water daily (30). Furthermore, populations living in rural Ethiopia are exposed to many risk factors that would negatively impact their physical function, including access to nutrition (31–33), healthcare, and sanitation (34–36). According to the 2016 DHS report, 25% of women living in rural Ethiopia were underweight (BMI < 18.5 ^kg^/_m_^2^), and 25% were anemic (30). Declines in physical function would significantly impact both necessary income-generating activities and activities needed for daily life.

Physical function is measured through physical tests, or validated survey questions focused on the respondents’ ability to complete tasks required for personal care and independent living (37). Several tests, including usual gait speed (UGS) and sit-to-stand tests (STS), do not require specialized training or resources, making them a viable option for measuring physical function in resource-limited settings. Gait speed strongly relates to pathophysiological mechanisms (38) and is considered a global marker of well-being (39). The STS requires a combination of muscle power and balance (40). It uses upper and lower-body musculature, requires continuous balance adjustments, and employs a motor pattern commonly used during ADLs (41). Both tests collect continuous outcome measures, which collect data on a gradient at the upper end of the functional spectrum, making them more useful in young and active populations (5,42,43).

Survey questions require less time and training to administer than direct tests, making them suitable for population studies. ADL surveys are one example of the validated survey questions used to measure physical function (44). They use population-specific questions to determine how well participants complete their ADLs (19,45). For example, surveys for elderly or infirmed populations often ask about participants’ ability to bathe, dress, and feed themselves (1,20,37,44).

Physical function is not well understood in rural Ethiopia because the physical function assessment tools have not been studied in this setting. The objectives of this study were to determine the feasibility, reliability, and validity of the UGS, STS, and ADLs survey to measure physical function in women living in rural highland Ethiopia. By understanding physical function in the region, future research could identify those needing additional healthcare and factors influencing health.

## Materials and Methods

### Study setting

Data were collected in the mountainous Central, Eastern, and South Zones of Tigray, Ethiopia. Most people in the rural highland regions have livelihoods based on rainfed agriculture. A short rainy season occurs between March and May, and the main rainy season occurs between June and October. During the main rainy season, more than 90 percent of the country’s total crop production occurs, with harvesting occurring between October and December (46). In addition, during the small rainy season, the poorest households in the regions receive additional support from a government social safety net called the Productive Safety Net Program (PSNP). The PSNP provides cash or food payments for work on community projects such as terracing land, building roads, water reclamation, and reforestation efforts (46,47).

Participants in this study belonged to households who had previously taken part in an evaluation of the PSNP (46). According to this evaluation, the villages included in this study were remote. The average travel time to the market was 85 minutes on foot. Only 1% of households had water piped into their dwelling; the average time to fetch water and return was 45 minutes. Female respondents spent most of their time conducting childcare, cooking/ eating, domestic activities, and personal care during the dry season. During the main rainy season, they spent most of their time on childcare, cooking/ eating, and food production activities (46).

### Study subjects

Participants in this study resided in households that had previously taken part in an evaluation of the PSNP (46). They were recruited based on the zones, counties, and villages where they lived. Within the zones, counties were excluded if the PSNP was inactive. Within each county, villages were excluded if they did not meet the criterion for a concurrently running study on physical exertion (48) which required two households that received benefits from the public works program and two that did not. All PSNP evaluation participants living in the selected villages were screened for eligibility. Prior studies that validated ADL questionnaires ranged in sample size from 30 (49,50) to over 400 people (51). Inclusion criteria included women aged 18-45 years who were not pregnant and were not planning on leaving the area during data collection. One hundred and seventy-eight women participated in the first round of data collection during the dry season, and 138 women participated in the second round of data collection during the main rainy season, the agricultural round. One hundred and twenty-seven women participated in data collection during both rounds; 51 only participated during the dry season round and 11 during the agricultural round.

### Data collection

Data collection took place over two four-week periods. The first four-week period, April-May 2019, coincided with the small rainy season when minimal agricultural activities occurred, and the PSNP was operational. The second, four-week period, September-October 2019, occurred during the main rainy season when households were engaged in farming activities such as preparing the land, planting, and weeding. The PSNP was non-operational.

Over eight days, participants were visited up to six times. Participants answered survey questions during three visits, including the ADL questions, and completed the physical function tests. During the first visit, relevant anthropometric information, including weight (Seca, Model 874 dr, Hamburg, Germany) and leg length, were also measured. Women wore light clothing while being weighed. Leg length was measured from the greater trochanter to the lateral malleolus using a standard measuring tape. All anthropometric measurements were measured twice, and the average of the two was used for analysis. Additional anthropometric, demographic, and village information including was collected during the PSNP evaluation (46).

### Physical function instruments

#### Sit-to-stand

A repeated STS test measures the time it takes a participant to rise from a seated position without using their hands. This task requires a combination of muscle power and balance (40). It uses upper and lower-body musculature, requires continuous balance adjustments, and employs a motor pattern commonly used in ADLs (41). There is no standardized protocol for this test, and several variations exist. For this study, we adapted the protocol used by Takai et al. (52), modifying the seated position and the number of tests to make them appropriate for this setting. The subjects started in a standing position, then sank into a low seated squat before standing again. They squatted seven times as quickly as possible. The bottoms of their feet remained in contact with the ground throughout the test. The low seated squat is a common position for adults in developing countries to adopt when sitting or resting. Participants crossed their arms at the wrists and held them against their chest to ensure they were unaided in rising.

A stopwatch recorded the time to the nearest 10^th^ of a second. The STS test was performed twice with an interval of 1 minute between the trials. The shorter of the two times was used for data analysis. A power index (P_sts_) was calculated using Equation 1. Previous studies have found that the power index is correlated with the maximum voluntary isometric knee extension force (52).

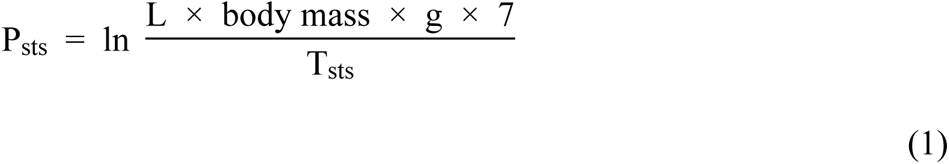

Where: P_sts_ is the power index from the test

L is the leg length measured from the greater trochanter to the lateral malleolus

Body mass is measured in kg to the nearest tenth

G is the acceleration of gravity (9.8 m/s^2^)

T_sts_ is the time it took to complete the test

#### Usual gait speed

During the UGS, participants were timed as they walked 20 meters at their usual pace. Enumerators laid out pre-measured lengths of rope on a flat surface, free of obstacles, on stable ground. The participants started three steps behind the start of the distance over which they were recorded walking and continued walking until they were three steps beyond the finishing point. The test was repeated three times, and the two closest times (out of three) were averaged and used for data analysis.

#### Activities of daily living survey

ADL surveys have been used to assess physical function since they were developed in 1963 (53). Multiple versions of the survey exist. Each version asks the study participants if they experienced difficulty completing routine activities such as getting in/out of bed, showering, and preparing food. The routine activities selected for this version of an ADL survey were based on pilot data from a “24-hour recall of time use” and “perceived energy exertion” survey conducted by the study team. Three ADLs were selected that almost all participants had engaged in and were not expected to vary much in difficulty throughout the year. The ADLs survey questions collected information on participants’ perceived difficulty (i) traveling to and from the market (ADL - Market), (ii) cleaning and housework (ADL - Housework), and (iii) preparing food (ADL - Food) due to the physical nature of the activity.

The participants ranked the level of difficulty encountered on an ordinal scale 1) experienced no difficulty, 2) experienced some difficulty, 3) needed assistance to complete the task, 4) avoided completing the task unless necessary, and 5) not responsible for the task. The responses to the ADL survey questions were collapsed into binary values whether the participant experienced no difficulty or any level of difficulty. Participants who were not responsible for a task were marked as “missing.” Additionally, a binary variable indicated whether a participant responded positively to experiencing difficulties with any ADLs (ADL – Any), and another variable captured the number of ADLs a participant reported difficulties (ADL – Number).

### Analysis

Feasibility was assessed based on the training and tools needed to administer the tests and qualitative reports from the enumerators on their ability to administer them. The reliability of each physical function test was evaluated using the Pearson correlation coefficient (ρ). The physical function tests from each visit were compared to the other visits with data. A coefficient between 0 to 0.20 was poor agreement, 0.21 to 0.40 was fair agreement, 0.41 to 0.60 was moderate/ acceptable agreement, 0.61 to 0.80 was substantial agreement, and 0.81 to 1.0 was near perfect agreement. These classifications have been used in other validation studies (54). Reliability for each ADL question was evaluated using Cohen’s kappa statistic (κ). The same cutoffs were used to assess agreement for the correlation coefficient (55).

Due to the low agreement between UGS and P_sts_ and low agreement of UGS between visits, only P_sts_ was used to assess the validity of the ADL survey questions. One visit per participant per round was randomly selected for analysis for the validity of the ADL survey. First, a univariate analysis was conducted to determine if the responses to the ADL questions were significantly associated with P_sts_. Then, the variables were regressed with the rounds collapsed and by round. In rounds where the p-value for an ADL question was less than 0.20, the ADL question was fit into a multivariate regress with P_sts_ as the dependent variable, and significant participant characteristics from that round were considered in the model. Significant participants characteristics included BMI, underweight (BMI < 18.5 kg/m2), having a partner live in the household, being food insecure as measured by food gap (46), self-reported short- or long-term illness, and residing in a household that benefits from the PSNP. The best fit modeled was determined using backward-stepwise regression by Bayesian information criterion (BIC).

### Ethical approval

This study received ethical approval from the IRB at Cornell University (1902008596).

## Results

Between the two rounds of data collection, participants were comparable in age, education, BMI, marital status, PSNP beneficiary status, and their self-reported long-term and short-term illnesses and injuries, **Table 1**. The mean age of the participants was 30 years, and the range was 18 to 45. The average BMI was 19.4 ^kg^/_m_^2^. However, more participants were underweight in the dry season round than in the agricultural (35.4% versus 31.6%). In addition, there was a seasonal difference in food security, with more participants reporting a food gap in the past six months during the dry season round (33.1%) than in the agricultural round (26.8).

**Table 1:** Study participant characteristics

| Characteristic | Dry season round | Agricultural round | P-value |
| --- | --- | --- | --- |
| **N** | **178** | **138** |  |
| Age (years) | 30 (6.6) | 30 (6.5) | 0.46 |
| Any education | 50.0% | 51.0% | 0.90 |
| BMI ($\text{kg}/\text{m}^2$) | 19.4 (2.1) | 19.5 (2.3) | 0.54 |
| Underweight (BMI < 18.5 $\text{kg}/\text{m}^2$) | 35.4% | 31.6% | 0.48 |
| Partner lives in household | 85.4% | 83.3% | 0.62 |
| Food insecure during the past 6 months | 33.1% | 26.8% | 0.23 |
| Long-term illness or injury | 15.7% | 14.5% | 0.76 |
| Short-term illness or injury | 24.7% | 26.8% | 0.67 |
| Benefits from PSNP | 47.2% | 44.9% | 0.69 |
| $P_{\text{sts}}$ | 9.9 (0.3) | 9.9 (0.3) | 0.89 |
| UGS | 8.7 (1.3) | 8.5 (1.4) | 0.26 |
| ADL - Market | 14.0% | 21.0% | 0.10 |
| ADL - Housework | 12.4% | 20.3% | 0.06 |
| ADL - Food | 12.4% | 21.0% | 0.04* |
| ADL - Any | 17.4% | 26.1% | 0.06 |
| ADL - Number |  |  |  |
| Zero | 82.6% | 73.9% | 0.06 |
| One | 5.1% | 6.5% | 0.58 |
| Two | 3.4% | 2.9% | 0.81 |
| Three | 9.0% | 16.7% | 0.04* |

The average values of the two physical function tests were similar between rounds. More participants reported difficulty with ADL - Any and each ADL during the agricultural round compared to the dry season round. There was only a statistically significant difference (p-value = 0.04) between those that reported difficulty preparing food for each round (12.4% vs 21.0%). During the dry season round, 82.6% of participants reported experiencing no difficulties with ADLs; this decreased to 73.9% in the agricultural round. In the dry season round, 9.0% of participants reported difficulties with all three ADLs. This increased to 16.7% of participants in the agricultural round (p-value = 0.04).

The enumerators reported that the STS was feasible to administer to the study population. Participants could complete the number of repetitions but reported that it was challenging and had started to fatigue. There were more reported difficulties in administering the UGS, particularly in finding a sufficient area to administer the tests. The UGS was not feasible in some households, making it an unsuitable test for use in rural Ethiopia.

The P_sts_ had a near-perfect agreement for each visit within a round, with a correlation coefficient greater than 0.80 (**Table 2**), except for visit 1 compared to visit 3 in the dry season round. There was substantial agreement between the values. Overall, UGS had a lower agreement, with most comparisons having moderate agreement. There was substantial agreement between visits 2 and 3 in the agricultural round (correlation coefficient = 0.62). There was nearly always poor agreement between the UGS and P_sts_ between each visit for both rounds.

**Table 2:** Reliability of physical function tests.

|  |  | Dry season round |  | Agricultural round |  |
| --- | --- | --- | --- | --- | --- |
| Test | Visit numbers | N | Pearson's ρ | N | Pearson's ρ |
| P<sub>sts</sub> |  |  |  |  |  |
|  | 1 vs 2 | 176 | 0.81**** | 126 | 0.88**** |
|  | 1 vs 3 | 147 | 0.78*** | 122 | 0.82**** |
|  | 2 vs 3 | 147 | 0.81**** | 129 | 0.84**** |
| UGS |  |  |  |  |  |
|  | 1 vs 2 | 176 | 0.54** | 124 | 0.59** |
|  | 1 vs 3 | 151 | 0.40* | 117 | 0.40* |
|  | 2 vs 3 | 152 | 0.47** | 121 | 0.62** |
| P<sub>sts</sub> vs |  |  |  |  |  |
| UGS |  |  |  |  |  |
|  | Visit 1 | 175 | -0.22* | 127 | -0.12 |
|  | Visit 2 | 176 | -0.10 | 132 | -0.05 |
|  | Visit 3 | 147 | -0.18 | 124 | -0.05 |

\* Indicates fair agreement, \*\* indicates moderate/ acceptable agreement, \*\*\*
indicates substantial agreement, \*\*\*\* indicated near-perfect agreement.

The responses to each ADL question were compared between visits in each round, **Table 3**. During the dry season round, there was a near-perfect agreement in the responses to the ADL questions between the visits (κ ranged from 0.78 to 0.97). During the agricultural round, there was either substantial or near-perfect agreement (κ ranged from 0.70 to 0.88).

**Table 3:**
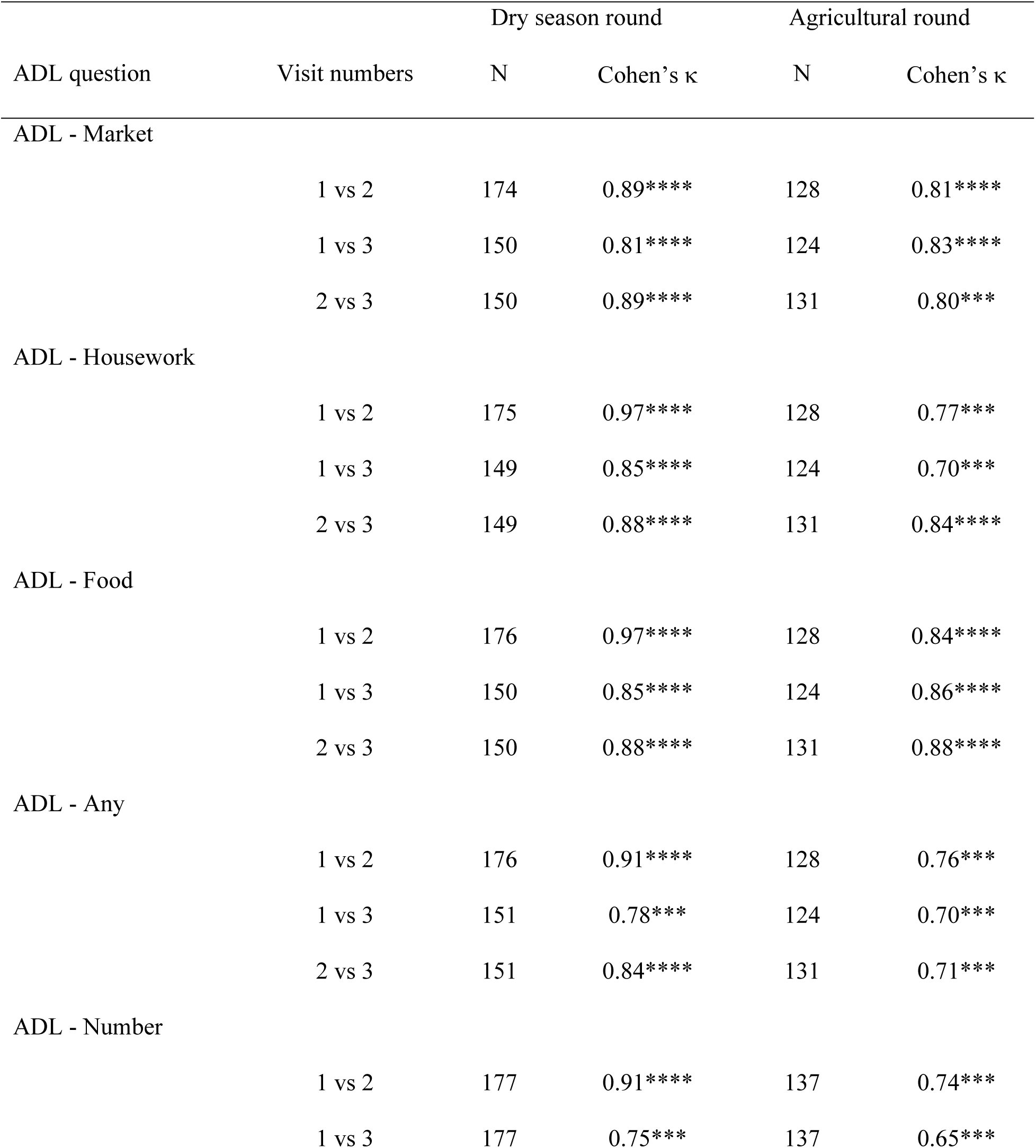
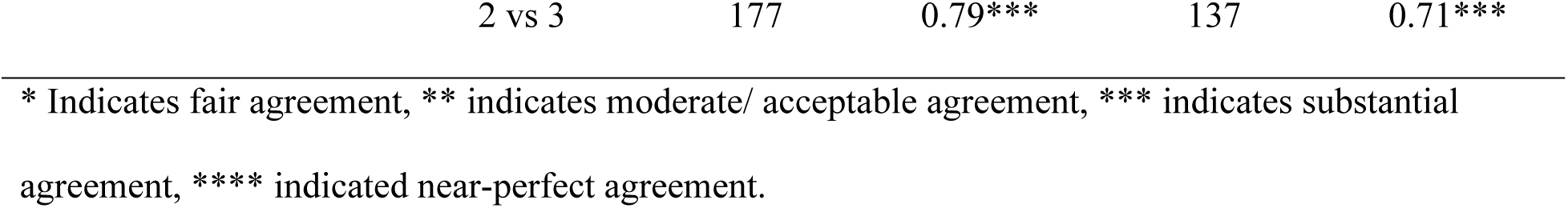
Correlation of ADL questions.

**Table 4** shows the association between Psts and participant characteristics and ADL responses with the rounds combine and by round. Age, BMI, underweight, and self-reported long-term illness or injury were consistently significantly associated with P_sts._ (p-value < 0.05). For every year increase in age, physical function as measured by the P_sts_ decreased. An increase in BMI was associated with improvements in physical function as measured by P_sts_, while those underweight had lower physical function as measured by P_sts_. Those that self-reported a long-term illness or injury had a lower physical function than those that did not. Among the ADL responses, none were significantly associated with P_sts_ when the rounds were combined. During the dry season round, ADL – Housework, ADL – Food, ADL – Any, and ADL – Number were significant at the α = 0.10. Only the ADL - Market was significant at the α = 0.10 level during the agricultural round.

**Table 4:** Association of P_sts_ with participant characteristics and ADL responses.

| Model parameter | All |  | Dry season round |  | Agricultural round |  |
| --- | --- | --- | --- | --- | --- | --- |
|  | Coefficien<br>t | p-value | Coefficien<br>t | p-value | Coefficien<br>t | p-value |
| N | 316 |  | 178 |  | 138 |  |
| Age | -0.01 | 0.01** | -0.01 | 0.06* | -0.01 | 0.04** |
| Any education | -0.02 | 0.46 | -0.01 | 0.88 | -0.04 | 0.30 |
| BMI (kg/m<sup>2</sup>) | 0.04 | <0.01*** | 0.05 | <0.01*** | 0.03 | <0.01*** |
| Underweight (BMI < 18.5 kg/m<sup>2</sup>) | -0.15 | <0.01*** | -0.18 | <0.01*** | -0.11 | 0.02** |
| Partner lives in household | 0.05 | 0.29 | 0.05 | 0.41 | 0.04 | 0.50 |
| Food insecure | -0.05 | 0.13* | -0.01 | 0.78 | -0.11 | 0.02** |
| Long term illness | -0.13 | 0.04** | -0.13 | 0.04** | -0.14 | 0.02** |
| Short term illness | 0.01 | 0.78 | 0.02 | 0.74 | -0.04 | 0.37 |
| Safety net beneficiary | 0.04 | 0.17* | 0.03 | 0.51 | 0.06 | 0.16* |
| ADL – Market | -0.01 | 0.82 | 0.08 | 0.21 | -0.10 | 0.07* |
| ADL – Housework | 0.02 | 0.59 | 0.12 | 0.09* | -0.06 | 0.29 |
| ADL – Food | 0.03 | 0.52 | 0.12 | 0.08* | -0.05 | 0.33 |
| ADL - Any | 0.03 | 0.53 | 0.12 | 0.08* | -0.05 | 0.33 |
| Number of ADLs | 0.01 | 0.74 | 0.04 | 0.09* | -0.03 | 0.17 |

The mean value of $\ln(P_{sts})$ for each round was 9.92 with a standard deviation of 0.30 for round 1 and 0.25 for round 2. \* Indicates statistical significance less than 0.20, qualifying for consideration in the multivariate model, \*\* Indicates statistical significance at <0.05, \*\*\* Indicates statistical significance <0.01

Multivariate models were constructed for each ADL question where the response was significant at α = 0.10, Table 5. In the dry season round, the ADL - Housework and ADL - Any were significant after controlling for BMI. The ADL - Food and ADL – Number were significant after controlling for BMI and self-reported long-term illness or injury. For the agricultural round, ADL-Market was significant after controlling for BMI. In the dry season round, all significant ADL variables had a positive coefficient, indicating that they were positively associated with P_sts_ and had improved physical function compared to those that responded that they had no difficulties. During the agricultural round, ADL - Market had a negative coefficient, indicating that those who reported difficulty had lower P_sts_ and lowered physical function than those who responded that they had no difficulties.

**Table 5:** Multivariate model ADLs and P_sts_.

| ADL question | ADL |  | BMI |  | Long-term illness or injury |  |
| --- | --- | --- | --- | --- | --- | --- |
|  | Coefficient | p-value | Coefficient | p-value | Coefficient | p-value |
| Dry season round |  |  |  |  |  |  |
| Clean | 0.13 | 0.04** | 0.05 | 0.02** |  |  |
| Food | 0.15 | 0.02** | 0.05 | <0.01*** | -0.12 | 0.04** |
| Any ADL | 0.12 | 0.03** | 0.05 | <0.01*** |  |  |
| Number of ADL | 0.05 | 0.03** | 0.05 | <0.01*** | -0.12 | 0.05** |
| Agricultural round |  |  |  |  |  |  |
| Travel | -0.10 | 0.04** | 0.03 | <0.01*** |  |  |

The mean value of $P_{sts}$ for each round was 9.92 with a standard deviation of 0.30 for round 1 and 0.25 for round 2. \* Indicates statistical significance less than 0.20, \*\* Indicates statistical significance at <0.05, \*\*\* Indicates statistical significance <0.01

## Discussion

This research found that P_sts_ (power index) is a feasible, reliable, and valid method to measure physical function in adult women living in rural highland Ethiopia. Measuring P_sts_ required very little training or materials, making it a low-cost, feasible option for research conducted in resource-constrained settings. Additionally, enumerators reported that the test was easy to administer and acceptable to the study participants. P_sts_ had high reliability with a near-perfect agreement between visits in each round. Additionally, P_sts_ was associated with age, BMI, and self-reported long-term illness or injury, indicating it is a valid measure of physical function. P_sts_ is a promising tool for measuring physical function in similar populations. UGS was neither feasible nor reliable, and it is not likely a good measure of physical function in adult women living in rural highland Ethiopia. The ADL survey was reliable, but it had inconclusive validity. The high reliability of the subjective scoring for each ADL among the rounds indicates that the participants understood the question and interpreted it consistently. More work is needed for it to be used to measure physical function in rural Ethiopia.

Strengths of this research include the careful attention to the development of the methodologies and the administration of the STS and UGS in a novel setting. Using P_sts_ as a measure of physical function is a strength of this paper. Previous work on physical function in LMICs used grip strength (27,29,56). To the authors’ knowledge, P_sts_ has not previously been used in rural regions of LMIC. P_sts_ is a better measure of physical function than grip strength because it requires muscle strength, mobility, balance, and coordination (40). In addition, P_sts_ is more likely to reflect the necessary activities to fulfill a study participant’s daily roles and responsibilities (41).

There were limitations in this paper, particularly around the development of the ADLs. The three activities were selected because they appeared in most responses in the time-use pilot survey. There was minimal seasonal variation in the frequency or difficulty of completing these activities. Unfortunately, logistical and time constraints prevented the piloting of these questions before the study. Questions added to the agricultural round of data collection found that these activities were the ones that were most likely to be altered when a participant was unwell. However, this was not reflected in that data. There is a lack of information to help us understand why the ADL questionnaire had limited validity or to aid in creating a new survey.

More research is needed to explore survey questions and activities that accurately characterize this population’s physical function. Additionally, research is required to understand the cause of the reported difficulties in completing ADLs. While the ADL questions may not detect decreased physical function, the responses should not be ignored or dismissed.

Future research that includes P_sts_ should examine several additional factors that impact physical function. This research’s nutritional status was limited to BMI, but it should include body composition. Physical function is both a cause and a consequence of lean body mass, making it an important consideration when evaluating physical function. Additionally, more detailed dietary intake and biomarkers such as hemoglobin should be measured. Diet quality, especially protein and micronutrient intake, impacts physical function (31–33). Physical activity, including type, intensity, and duration, should be included in future research. Medical history was limited to self-reported long or short-term illness or injury in this research, but an in-depth medical history would be valuable to future research, including the type, timing, treatment, and health-seeking behaviors around illnesses and injuries. Finally, research regarding physical function needs more frequent data points over longer periods. There are seasonal variations in many factors that influence physical function, including diet (57), physical activity (46), and exposure to diseases (58). More data points will help determine how quickly physical function changes in women in rural Ethiopia in response to different factors and if there are long-term changes.

The STS was piloted among the enumerators before the start of the study. There was not a consistent chair height among the households of the study participants, eliminating the possibility of using a chair for the seated position. Like other cultures, people in rural Ethiopia frequently sit in a deep squatted position while resting or working near the ground (e.g., farming activities such as planting and weeding or domestic work such as cooking or laundry). As such, the study team determined that it would be an appropriate option for the seated position of the STS, especially since the P_sts_ variable took leg length into account. The enumerators determined that seven was the ideal number of repetitions. It was challenging enough to reflect variations in physical function, but participants could complete it.

## Conclusions

Measuring physical function and identifying those experiencing difficulties completing their ADLS is essential in all vulnerable populations, including women in rural Ethiopia. ADLs in rural highland Ethiopia are physically demanding. Taking care of oneself, family, and household, and many income-generating activities require significant manual labor and traveling significant distances on foot. Therefore, decreases in physical function could affect the well-being of one’s family by impacting household income, diet quality, and health through water, sanitation, and hygiene.

A feasible and reliable measure of physical function is vital in addressing poverty and malnutrition. It would allow healthcare providers to identify those requiring additional medical care and researchers to identify factors that influence physical function, including nutritional, behavioral, and environmental factors. This research has demonstrated that the P_sts_ using a timed seven-repetition stand to seated squat is a reliable measure of physical function in this setting.

## Data Availability

All data is available https://doi.org/10.7910/DVN/QEJGEO

## Acknowledgements

We would like to thank our field research staff in Tigray, Ethiopia, especially Equbay Gebrehiwet, for all their work during data collection. Additionally, we are thankful for the support provided by the Ethiopian Strategic Support of the International Food Policy Research Institute. We would like to thank Drs Kathleen M. Rasmussen and James Booth for their support.

## Notes

### Competing Interest Statement

The authors have declared no competing interest.

### Funding Statement

JG was supported by the National Institutes of Health under award T32-DK007158 while conducting this research. https://www.niddk.nih.gov/ NO JH does not have any disclosures to report. ATM was funded by the International Council on Amino Acid Science and the U.S. Highbush Blueberry Council. There aren’t grant numbers associated with these. https://icaas-npo.org/english/index.html https://ushbc.blueberry.org/ NO: The funders had no role in study design, data collection and analysis, decision to publish, or preparation of the manuscript

### Author Declarations

Cornell University Institutional Review Board for Human Participants provided approval of research. Protocol ID#: 1902008596 Written consent was obtained

